# Unusual SARS-CoV-2 intra-host diversity reveals lineages superinfection

**DOI:** 10.1101/2021.09.18.21263755

**Authors:** Filipe Zimmer Dezordi, Paola Cristina Resende, Felipe Gomes Naveca, Valdinete Alves do Nascimento, Victor Costa de Souza, Anna Carolina Dias Paixão, Luciana Appolinario, Renata Serrano Lopes, Ana Carolina da Fonseca Mendonça, Alice Sampaio Barreto da Rocha, Taina Moreira Martins Venas, Elisa Cavalcante Pereira, Richard Steiner Salvato, Tatiana Schäffer Gregianini, Leticia Garay Martins, Felicidade Mota Pereira, Darcita Buerger Rovaris, Sandra Bianchini Fernandes, Rodrigo Ribeiro-Rodrigues, Thais Oliveira Costa, Joaquim Cesar Sousa, Fabio Miyajima, Edson Delatorre, Tiago Gräf, Gonzalo Bello, Marilda Mendonça Siqueira, Gabriel da Luz Wallau, on behalf of Fiocruz COVID-19 Genomic Surveillance Network

**Affiliations:** Departamento de Entomologia, Instituto Aggeu Magalhães (IAM), FIOCRUZ-Pernambuco, Recife, Pernambuco, Brasil; Núcleo de Bioinformática (NBI), Instituto Aggeu Magalhães (IAM), FIOCRUZ-Pernambuco, Recife, Pernambuco, Brasil; Laboratório de Ecologia de Doenças Transmissíveis na Amazônia (EDTA), Instituto Leônidas e Maria Deane, FIOCRUZ-Amazonas, Manaus, Amazonas, Brasil; Laboratory of Respiratory Viruses and Measles (LVRS), Instituto Oswaldo Cruz, FIOCRUZ-Rio de Janeiro, Rio de Janeiro, Rio de Janeiro, Brasil; Laboratório Central de Saúde Pública, Centro Estadual de Vigilância em Saúde da Secretaria de Saúde do Estado do Rio Grande do Sul (LACEN/CEVS/SES-RS), Porto Alegre, Rio Grande do Sul, Brasil; Centro Estadual de Vigilância em Saúde da Secretaria de Saúde do Estado do Rio Grande do Sul, Porto Alegre, Rio Grande do Sul, Brasil; Laboratório Central de Saúde Pública do Estado da Bahia (LACEN-BA), Salvador, Bahia, Brasil; Laboratório Central de Saúde Pública do Estado de Santa Catarina (LACEN-SC), Florianópolis, Santa Catarina, Brasil; Laboratório Central de Saúde Pública do Estado do Espírito Santo (LACEN-ES), Vitória, Espírito Santo, Brasil; Analytical Competence Molecular Epidemiology Laboratory (ACME), FIOCRUZ-Ceará, Fortaleza, Ceará,Brasil; Departamento de Biologia. Centro de Ciências Exatas, Naturais e da Saúde, Universidade Federal do Espírito Santo, Espírito Santo, Alegre, Brasil; Plataforma de Vigilância Molecular, Instituto Gonçalo Moniz, FIOCRUZ-Bahia, Salvador, Bahia, Brasil; Laboratório de AIDS e Imunologia Molecular, Instituto Oswaldo Cruz, FIOCRUZ-Rio de Janeiro, Rio de Janeiro, Rio de Janeiro, Brasil

**Keywords:** codetection, coinfection, COVID-19, genomics

## Abstract

The SARS-CoV-2 has infected almost 200 million people worldwide by July 2021 and the pandemic has been characterized by infection waves of viral lineages showing distinct fitness profiles. The simultaneous infection of a single individual by two distinct SARS-CoV-2 lineages provides a window of opportunity for viral recombination and the emergence of new lineages with differential phenotype. Several hundred SARS-CoV-2 lineages are currently well characterized but two main factors have precluded major coinfection/codetection analysis thus far: i) the low diversity of SARS-CoV-2 lineages during the first year of the pandemic which limited the identification of lineage defining mutations necessary to distinguish coinfecting viral lineages; and the ii) limited availability of raw sequencing data where abundance and distribution of intrasample/intrahost variability can be accessed. Here, we have put together a large sequencing dataset from Brazilian samples covering a period of 18 May 2020 to 30 April 2021 and probed it for unexpected patterns of high intrasample/intrahost variability. It enabled us to detect nine cases of SARS-CoV-2 coinfection with well characterized lineage-defining mutations. In addition, we matched these SARS-CoV-2 coinfections with spatio-temporal epidemiological data confirming their plausibility with the co-circulating lineages at the timeframe investigated. These coinfections represent around 0.61% of all samples investigated. Although our data suggests that coinfection with distinct SARS-CoV-2 lineages is a rare phenomenon, it is likely an underestimation and coinfection rates warrants further investigation.

**DATA SUMMARY:** The raw fastq data of codetection cases are deposited on gisaid.org and correlated to gisaid codes: EPI_ISL_1068258, EPI_ISL_2491769, EPI_ISL_2491781, EPI_ISL_2645599, EPI_ISL_2661789, EPI_ISL_2661931, EPI_ISL_2677092, EPI_ISL_2777552, EPI_ISL_3869215. Supplementary data are available on https://doi.org/10.6084/m9.figshare.16570602.v1. The workflow code used in this study is publicly available on: https://github.com/dezordi/IAM_SARSCOV2.

## INTRODUCTION

The SARS-CoV-2, the etiological agent of the COVID-19 pandemic, has a relatively low mutation rate compared to other RNA viruses ^1^, and most viral lineages are normally defined by only a few synapomorphic SNPs (*n* < 10) ^2^. However, the pervasiveness of SARS-CoV-2 infections during the COVID-19 pandemic provided substantial opportunities for the virus to explore the fitness landscape through single nucleotide substitutions and/or indels, giving birth to a range of more transmissible variants of concern (VOCs). These lineages are characterized by an unusual pattern of lineage-defining SNPs along the genome (*n* > 15)^3,4,5^.

Coinfection is defined as a single cell/host infection by more than one virus lineage simultaneously. Despite a rare phenomenon, it may provide opportunity for genetic recombination, an event known to occur in viruses of the *Coronaviridae* family ^6,7^. Recombinant viruses may, in turn, trigger the emergence of new lineages with enhanced biological properties, including the capacity to infect new hosts (expansion of viral host range) ^8–11^. The frequency of coinfected patients and its role to promote recombination-driven SARS-CoV-2 evolution and the emergence of SARS-CoV-2 lineages is still poorly understood. The low variability found in SARS-CoV-2 lineages and the few well-defined lineage-specific SNPs until the second half of 2020 probably hindered the identification of coinfection and recombination events of SARS-CoV-2 lineages so far. In contrast the emergence of VOCs lineages carrying a substantial number of additional SNPs may provide enough markers to currently detect these events. A number of coinfection cases were reported for SARS-CoV-2, including lineages B.1.1.28/B.1.1.33 and B.1.1.91/B.1.1.28 ^12^ and several variants of interest (VOIs) and VOCs^13^. Moreover, putative coinfections were indirectly inferred from North America and Europe patients by detecting recombinant genomes ^14,15^.

In this study, we assessed amplicon sequencing reads of 2,263 SARS-CoV-2 samples from Brazilian patients generated by the Fiocruz Genomic Surveillance Network. We identified nine coinfection cases through the identification of an unusual pattern of intrasample/intrahost single nucleotide variation (iSNV) and phylogenetic reconstruction of alternative SARS-CoV-2 genomes generated by well supported iSNVs. Moreover, epidemiological trends of circulating lineages in each Brazilian state supported that the SARS-CoV-2 variant of interest (VOI) and VOCs lineages found in these coinfected samples were also co-circulating at the time of sampling, thus providing further plausibility for our findings.

## METHODS

### SARS-CoV-2 sequences and ethical aspects

The sequencing data was obtained through the genomic survey of SARS-CoV-2 positives samples sequenced by Fiocruz COV-19 Genomic Surveillance Network between 18 May 2020 and 30 April 2021. The SARS-CoV-2 genomes were recovered using previously described Illumina protocols ^16–18^ (**Table S1)**. The frequency of lineages by Brazilian states was evaluated using data recovered from GISAID (gisaird.org) on 23 July 2021. All samples used in this work are approved by the Ethics Committee of the Aggeu Magalhaes Institute Ethical Committee—CAAE 32333120.4.0000.5190, the Ethics Committee of Amazonas State University (no. 25430719.6.0000.5016), the Ethics Committee of FIOCRUZ-IOC (68118417.6.0000.5248) and the Brazilian Ministry of Health SISGEN (A1767C3).

### Genome assembly and intrahost variant analysis

The Fastq reads were submitted in an in house workflow available at https://github.com/dezordi/IAM_SARSCOV2 that performs the following steps: The remotion of duplicated reads, adapters and read extremities with less than 20 of phred score quality with the fastp tool ^19^; A genome assembly guided by reference was performed with BWA ^20^ mapping reads against the SARS-CoV-2 Wuhan reference genome (NC_045512.2); The consensus genomes were generated with samtools mpileup ^21^ and iVar ^22^, using a threshold quality score of 30 and calling SNPs and indels present as major allele frequencies; After the consensus generation, the bam-readcount tool ^23^ was used to retrieve the proportion of each base (A, C, T, G) present in each position of the bam file and a in house python script (intrahost.py) was used to identify position with putative intra-host variants related to minor allele frequencies following specific rules: The minor variant (MinV) should represent at least 5% of total position depth, and the MinV should have present in reads of both senses (at least 5% in each sense) with at least 100 reads of depth. At the end of the workflow, two consensus genomes were generated, one with the MajV (the nucleotide present in major allele frequency) in each genomic position showing well supported alternative nucleotides, and another with the MinV (the nucleotide present in the lower allele frequency) at that same positions; The consensus genomes were submitted to PangoLineage tool v1.1.23 and pangoLEARN update at 28 May 2021 ^24^ and to Nextclade ^25^ tools. Only genomes with more than 95% coverage breadth and 100 reads of average coverage depth (**Table S2**) were considered.

For all samples in which alternative genomes where assigned to two different pango lineages were manually curated with Interactive Genomic Viewer ^26^, indels related to intrahost variants into specific genomes that change the coding frame were discarded (insertions and deletions of one or two nucleotides). MajV and MinV genomic versions were generated based on the reasoning that each may represent a different SARS-CoV-2 lineage. If so, then each consensus should cluster within a cognate lineage in the phylogenetic analysis characterizing a codetection. Additional evidence of codetection was searched on the raw sequence reads: I - if the proportion of reads supporting lineage-specific defining SNPs are similar it suggests codetection while if the proportion is drastically different the variability is likely derived from de novo intrahost variability; II - if SNPs and/or intrahost variants are restricted to some specific SARS-CoV-2 genomic region it likely indicates a recombination event. Otherwise, if intrahost variants are distributed along the entire SARS-CoV-2 genome it is likely to be derived from the codetection of different SARS-CoV-2 genomes in the same sample.

### Phylogenetic Analysis

A reference alignment was created using MAFFT ^27^ with the 6,167 genomes, which represents the genomes present in the nextstrain ^28^ phylogeny with less than 5% of N content on 24th May 2021 and Brazilian genomes obtained through a cd-hit-est ^29^ clusterization of genomes present on GISAID at 16th March 2021 with high-quality, with more than 99.8% sequence identity and from the same Brazilian state. The reference alignment was edited to mask UTR regions and to maintain the indel regions. The 34 MajV and MinV consensus genomes were aligned to reference alignment with MAFFT add, and we performed a Maximum-Likelihood phylogenetic analysis with IQtree2 ^30^ using the aLRT strategy and the GTR+F+R5 model. The PANGO lineages were evaluated with pangolin and used to annotate the tree with iTOL ^31^.

## RESULTS AND DISCUSSION

Our initial analysis revealed that 1,462 out of 2,263 genomes had enough sequencing breadth and depth to be able to consistently detect and characterize the viral genomic variability at the sequencing reads level. 1,150 out of 1,462 SARS-CoV-2 positive samples investigated showed at least one genomic site with supported intra-host variability, that is, at least one genomic position with more than 100 reads supporting a minimum of two alternative nucleotides. Those samples showed an average coverage depth of 1817.46 (stdev = 908.59) and an average coverage breath supported by at least 100 reads of 99.66 (stdev = 1.10) (**Table S2**). In addition, we estimated a mean of 2.57 genomic sites showing intra-host variants (**Table S3**). Major and Minor consensus sequences were generated for all samples bearing well supported alternative nucleotides. These alternative consensus genomes, representing the viral genome variability found in each sample, were then assessed for lineage assignment using the PangoLineage tool. If the same lineage was recovered for both genomes, this represents that the Major and Minor variants did not differ in relation to lineage-defining SNPs and that the variability observed likely resulted from de novo intra-host variants that emerged during viral replication. Conversely, if Major and Minor genomic variants were assigned to different lineages, the intra-host variability observed is more likely derived from a codetection event. We detected 16 instances in which Major and Minor variants were assigned to distinct lineages (intra-host sites: mean = 24, stdev = 9.75), including former Variants for Further Monitoring (VFM) N.9 and P.2 as well as the high circulating VOC P.1 (**Table S4**). To further evaluate the lineage assigned by PangoLineage, we performed a phylogenetic analysis of representative lineages including both Major and Minor genomes, where true codetection is supported if each genome is placed into a different well supported clade/lineage. Nine alternative genomes were confidently repositioned into distinct lineages (**Figure 1**, red arrows, mean MAFs sites 30.44, stdev = 6.63), while the remaining alternative genomes remained in the same lineage (**Figure 1**, black arrows, mean MAFs 23.06, stdev = 10.54). Intra-host variants sites found showed several lineage defining SNPs spread across the whole SARS-CoV-2 genome, and the sequencing reads depth was roughly similar throughout the genome (**Figure 2A, Table S5**).

**Figure1.**
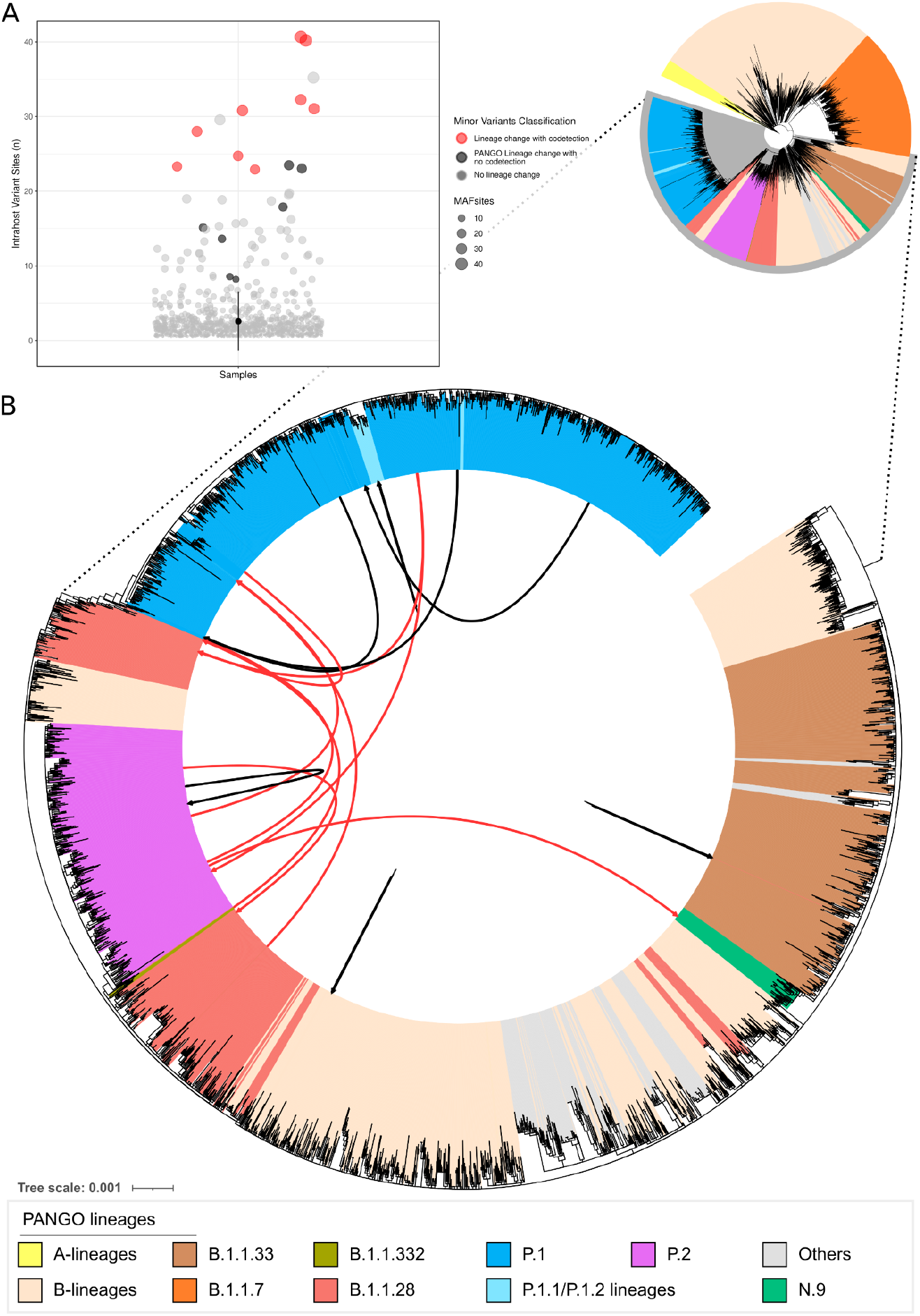
SARS-CoV-2 samples with MAFs. **A** - Dot Plot with number of MAFs per sample; **B** - Maximum likelihood phylogenetic tree. Others: R, S, U, L, D, C lineages. Red arrows represent samples with alternative genomes showing lineage change while black arrows indicate alternative samples with no lineage change.

**Figure 2.**
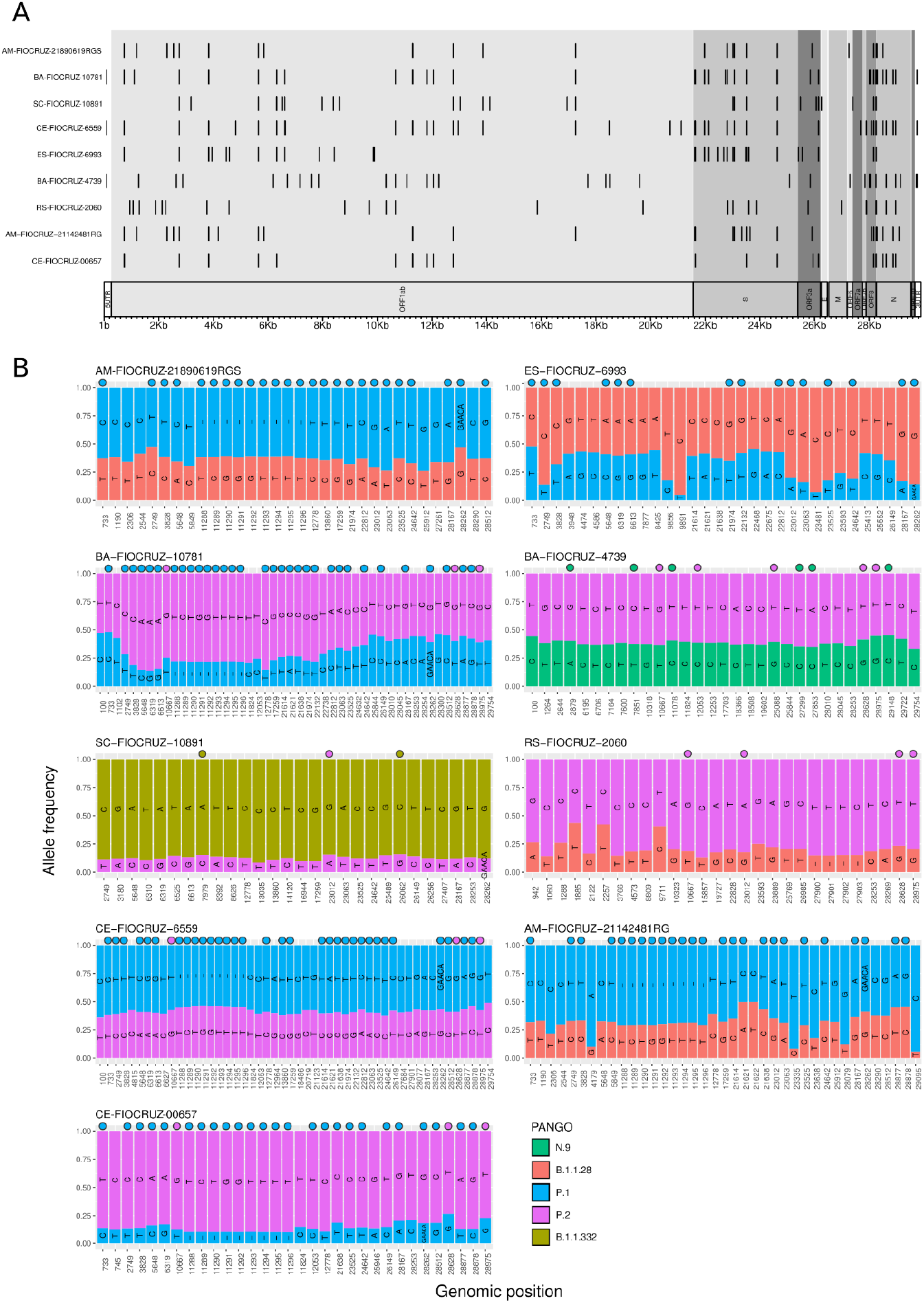
Major and minor allele frequencies of samples with codetection of different SARS-CoV-2 lineages. **A**. Karyoploter with MAFs sites across the SARS-CoV-2 genome. **B**. MAFs sites with read depth frequency supporting major and minor nucleotides. Defining SNPs based on data of outbreak.info update on 24 July 2021, are indicated with a circle. Karyoplots depicting MAF sites sequencing depth can be accessed on **Supplementary File 1**, and raw depth values can be accessed in **Table S5**.

Seven out of nine putative coinfection events involve the VOC Gamma (P.1 lineage) (**Table 1**). In four cases, P.1/Gamma lineage represented the major genomic variant, while in the remaining three cases, it corresponded to the minor variant. The large proportion of codetection events with P.1/Gamma is likely a result of the higher number of lineage-defining SNPs characteristic of this lineage that facilitate the distinction between coinfecting SARS-CoV-2 lineages. As more distinct lineages, bearing many lineage-defining SNPs, coinfect the same host, it becomes increasingly more likely to objectively distinguish the coinfecting lineages through the reconstruction of alternative intrasample viral genomes.

**Table 1.** Summary of coinfection events

| Sample | State | City | MAF sites | cov* | depth** | Lineages<sup>1</sup> | Collection Date | 1st MajV<sup>2</sup> lineage available on GISAID | 1st MinV<sup>2</sup> lineage available on GISAID |
| --- | --- | --- | --- | --- | --- | --- | --- | --- | --- |
| CE-FIOCRUZ-00657 | CE | Fortaleza | 23 | 98.95 | 762.04 | P.2/P.1 | 2021-01-20 | 2020-11-20/2020-04-15 | 2021-01-07/2021-01-07 |
| AM-FIOCRUZ-21142481RG | AM | Manaus | 31 | 98.9 | 1105.46 | P.1/B.1.1.28 | 2021-01-13 | 2020-12-03/2020-12-03 | 2020-04-13/2020-04-13 |
| RS-FIOCRUZ-2060 | RS | Canoas | 25 | 99.69 | 3509.92 | P.2/B.1.1.28 | 2021-01-07 | 2021-01-26/2020-08-31 | 2020-05-21/2020-03-16 |
| BA-FIOCRUZ-4739 | BA | Salvador | 31 | 99.73 | 5354.25 | P.2/N.9 | 2021-01-08 | 2020-10-26/2020-06-26 | 2020-12-10/2020-11-12 |
| ES-FIOCRUZ-6993 | ES | Aracruz | 32 | 99.67 | 2222.7 | B.1.1.28/P.1 | 2021-01-09 | 2020-10-13/2020-10-13 | 2021-04-09/2021-01-22 |
| CE-FIOCRUZ-6559 | CE | Fortaleza | 41 | 99.85 | 4090.76 | P.1/P.2 | 2021-01-24 | 2021-01-07/2021-01-07 | 2020-11-20/2020-04-15 |
| SC-FIOCRUZ-10891 | SC | Porto Belo | 28 | 98.72 | 2153.25 | B.1.1.332/B.1.1.28 | 2021-02-22 | 2021-02-22/2021-02-22 | 2020-03-18/2020-03-18 |
| BA-FIOCRUZ-10781 | BA | Salvador | 40 | 99.73 | 1996.63 | P.2/P.1 | 2021-02-10 | 2020-10-26/2020-06-26 | 2020-12-27/2020-12-27 |
| AM-FIOCRUZ-21890619RGS | AM | Manaus | 23 | 96.59 | 1018.89 | P.1/B.1.1.28 | 2021-01-13 | 2020-12-03/2020-12-03 | 2020-04-13/2020-04-13 |

<sup>1</sup>MajV/MinV Pango lineages supported into the phylogeny. <sup>2</sup>Date of the first genome deposited on GISAID of each variant into the specific municipality/state, updated on 26 July 2021. \* Coverage breadth supported by 100 reads; \*\* Average coverage depth. AM: Amazonas; BA: Bahia; ES: Espírito Santo; CE: Ceará; RS: Rio Grande do Sul; SC: Santa Catarina.

In order to assess if codetection could be a result of sample contamination we reassessed sample AM-FIOCRUZ-21142481RG from RNA extraction, library preparation and sequencing. We confirmed the intrahost variability for 25 out of 31 sites present in the first sequencing run (**Table S6**). Moreover, lineage assignment, phylogenetic reconstruction and the detection of SNP defining mutations confirmed the codetection status of that sample (**Table S4**).

The plausibility of the codetection events may be further vindicated by the fact of all major and minor alternative genomes lineages identified by our study were recognized co-circulating variants in their source of collection, overlapping in time and space, as well as matching with SARS-CoV-2 lineage information form their respective geographical states (**Figure 3**). The Major consensus genome corresponding to the predominant lineage circulating in Rio Grande do Sul, Bahia, and Amazonas states were recovered in our analysis. On the other hand, in Santa Catarina, both lineages involved in codetection were present at lower frequency than the dominant lineages at the same location and period of sampling. Only one event of VOC circulation without notification may have occurred in respect to a sample from Aracruz city, Espirito Santo state, where the VOC Gamma identified was in fact the Minor Variant genomic variant from a sample collected on 9th January 2021. Interestingly, from a direct search from GISAID, the earliest record of the VOC Gamma from Aracruz were from 4^th^ April 2021 and 22^nd^ January 2021 in the Espirito Santo state (**Table 1**), whilst the genomic sequence of this specimen deposited on GISAID under EPI_ISL_2645599 code confirmed that the deposit of the Major consensus genome reference to the B.1.1.28 lineage.

**Figure 3.**
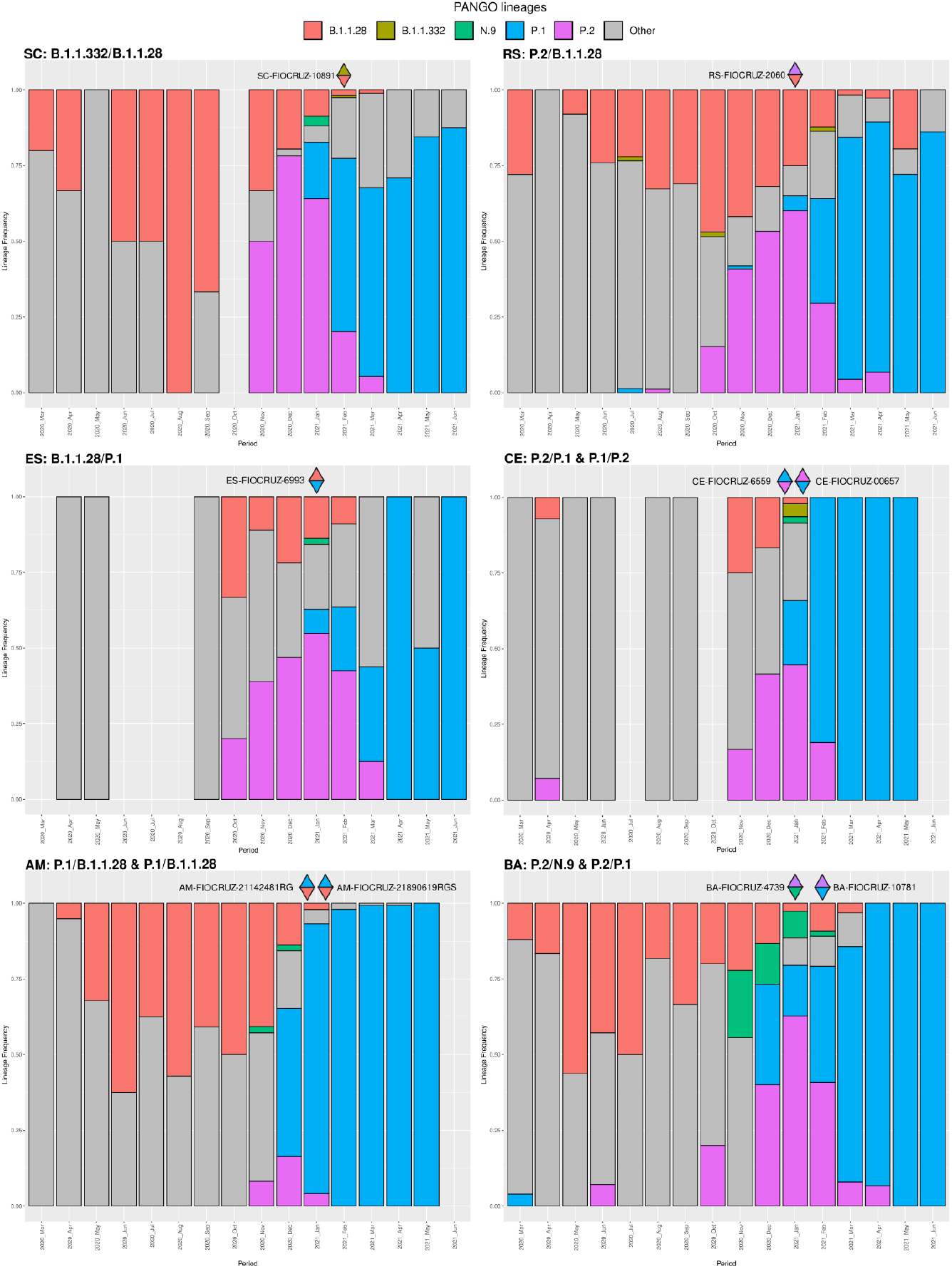
SARS-CoV-2 lineage proportion through time in different Brazilian states with codetection cases. Data were recovered from GISAID on 23 July 2021, raw data can be accessed in **Table S7**. Upper triangles colored with the lineage of major consensus genomes and lower triangles with minor consensus genomes lineages.

This study reports that codetection/coinfection events occurred at a low rate in Brazil (0.61% - 9 samples from 1462). This is certainly an underestimation due to the limitation of detecting true coinfection events of earlier low diverging SARS-CoV-2 lineages that dominated the first year of the pandemic. Despite that, considering the lower bound of recorded SARS-CoV-2 cases worldwide until July 2021 were around 190 million (https://coronavirus.jhu.edu/map.html), we can infer that at least 1,1 million patients have been coinfected across the world, which in turn provides a substantial window of opportunity for SARS-CoV-2 recombination events. Moreover, this estimate is certainly downwardly biased because the number of asymptomatic infections is largely not accounted for.

## Conclusions

In line with other studies, we showed that SARS-CoV-2 has an apparent low intrahost variability overall. Our in-depth analysis revealed at least nine codetection events which are corroborated by epidemiological data from co-circulating lineages in different Brazilian states. Moreover, the lineages identified revealed the early emergence of cryptically circulating lineages not detected by consensus genomes alone. The large number of genomic sites carrying alternative nucleotides in codetection events may generate artificial hybrid consensus genomes, therefore a careful inspection of consensus sequences against sequencing reads polymorphisms is warranted to generate robust consensus sequences. Considering the large case numbers of SARS-CoV-2 infections worldwide and that coinfection are more likely to happen in high transmission settings, all efforts should be placed to limit SARS-CoV-2 transmission hence reducing the likelihood of emergence of novel recombinant hybrid lineages with altered phenotype.

## Data Availability

The raw fastq data of codetection cases are deposited on gisaid.org and correlated to gisaid codes: EPI_ISL_1068258, EPI_ISL_2491769, EPI_ISL_2491781, EPI_ISL_2645599, EPI_ISL_2661789, EPI_ISL_2661931, EPI_ISL_2677092, EPI_ISL_2777552, EPI_ISL_3869215. Supplementary data are available on https://doi.org/10.6084/m9.figshare.16570602.v1. The workflow code used in this study is publicly available on: https://github.com/dezordi/IAM_SARSCOV2.

https://github.com/dezordi/IAM_SARSCOV2

## AUTHOR STATEMENTS

### Authors and Contributors

Conceptualisation: FZD and GLW. Methodology: FZD and GLW. Formal Analysis: FZD. Investigation: FZD, PCR, FGN, VAN, VCS., ACDP, LA, RSL, ACFM, ASBR, TMMV, ECP, RSS., TSG., LGM, FMP., DBR, SBF., RR, TOC, JCS, FM, ED, TG, GB, MMS. Resources: PCR, FGN, FM, ED, TG, GB, MMS, GLW. Writing: FZD, PCR, FGN, VAN, VCS., ACDP, LA, RSL, ACFM, ASBR, TMMV, ECP, RSS., TSG., LGM, FMP., DBR, SBF., RR, TOC, JCS, FM, ED, TG, GB, MMS, GLW.

### Conflicts of interest

The authors declare that there are no conflicts of interest.

### Funding information

Financial support was provided by FAPEAM (PCTIEmergeSaude/AM call 005/2020 and Rede Genômica de Vigilância em Saúde - REGESAM); Ministério da Ciência, Tecnologia, Inovações e Comunicações/Conselho Nacional de Desenvolvimento Científico e Tecnológico - CNPq/Ministério da Saúde - MS/FNDCT/SCTIE/Decit (grants 402457/2020-9 and 403276/2020-9); Inova Fiocruz/Fundação Oswaldo Cruz (Grants VPPCB-007-FIO-18-2-30 and VPPCB-005-FIO-20-2-87) and INCT-FCx (465259/2014-6). This work was also supported by the Pan American Health Organization, Brazil Country Office. FGN, GLW and GB are supported by the CNPq through their productivity research fellowships (306146/2017-7, 303902/2019, 302317/2017-1, respectively).

## Acknowledgments

We thank the Fiocruz COVID-19 Genomic Surveillance Network for sharing this large dataset and embracing such collaborative work. We also thank all the researchers around the world that are working and generating data of SARS-CoV-2 in those difficult times. The acknowledgment info of all SARS-CoV-2 genomes from GISAID and used in this work are present in **Supplementary File 2**.

## Notes

### Competing Interest Statement

The authors have declared no competing interest.

### Funding Statement

Financial support was provided by FAPEAM (PCTIEmergeSaude/AM call 005/2020 and Rede Genomica de Vigilancia em Saude - REGESAM); Ministerio da Ciencia, Tecnologia, Inovacoes e Comunicacoes/Conselho Nacional de Desenvolvimento Cientifico e Tecnologico - CNPq/Ministerio da Saude - MS/FNDCT/SCTIE/Decit (grants 402457/2020-9 and 403276/2020-9); Inova Fiocruz/Fundacao Oswaldo Cruz (Grants VPPCB-007-FIO-18-2-30 and VPPCB-005- FIO-20-2-87) and INCT-FCx (465259/2014-6). This work was also supported by the Pan American Health Organization, Brazil Country Office. FGN, GLW and GB are supported by the CNPq through their productivity research fellowships (306146/2017-7, 303902/2019, 302317/2017-1, respectively).

### Author Declarations

The study was approved by the Aggeu Magalhaes Institute Ethical Committee - CAAE 32333120.4.0000.5190, FIOCRUZ-IOC Ethics Committee (68118417.6.0000.5248 and CAAE 32333120.4.0000.5190), the Brazilian Ministry of Health SISGEN (A1767C3). Moreover, it was approved by the Ethics Committee of Amazonas State University (no. 25430719.6.0000.5016), which waived signed informed consent

